# High levels of common cold coronavirus antibodies in convalescent plasma are associated with improved survival in COVID-19 patients

**DOI:** 10.1101/2021.03.08.21252775

**Authors:** Uri Greenbaum, Kimberly Klein, Fernando Martinez, Juhee Song, Peter F. Thall, Jeremy L. Ramdial, Cristina Knape, Fleur M. Aung, Jamie Scroggins, Adriana Knopfelmacher, Victor Mulanovich, Jovan Borjan, Javier Adachi, Mayoora Muthu, Cerena Leung, Mayrin Correa Medina, Richard Champlin, Amanda Olson, Amin Alousi, Katayoun Rezvani, Elizabeth J. Shpall

## Abstract

**Background:** COVID-19 Convalescent plasma (CCP) is safe and effective, particularly if given at an early stage of the disease. Our study aimed to identify an association between survival and specific antibodies found in CCP.

**Patients and Methods:** Patients ≥18 years of age who were hospitalized with moderate to severe COVID-19 infection and received CCP at the MD Anderson Cancer Center between 4/30/2020 and 8/20/2020 were included in the study. We quantified the levels of anti-SARS-CoV-2 antibodies, as well as antibodies against antigens of other coronavirus strains, in the CCP units and compared antibody levels with patient outcomes. For each antibody, a Bayesian exponential survival time regression model including prognostic variables was fit, and the posterior probability of a beneficial effect (PBE) of higher antibody level on survival time was computed.

**Results:** CCP was administered to 44 cancer patients. The median age was 60 years (range 37-84) and 19 (43%) were female. Twelve patients (27%) died of COVID-19-related complications. Higher levels of two non-SARS-CoV-2-specific antibodies, anti-HCoV-OC43 spike IgG and anti-HCoV-HKU1 spike IgG, had PBE = 1.00, and 4 SARS-CoV-2-specific antibodies had PBEs between 0.90 and 0.95. Other factors associated with better survival were shorter time to CCP administration, younger age, and female sex.

**Conclusions:** Common cold coronavirus spike IgG antibodies anti-HCoV-OC43 and anti-HCoV-HKU1 may target a common domain for SARS-CoV-2 and other coronaviruses. They provide a promising therapeutic target for monoclonal antibody production.

## Introduction

Few treatment options for COVID-19 have proven effective in robust clinical trials, and while vaccines are now available for prevention of severe infections, more effective therapeutic agents are still an unmet need. This is especially true for immunocompromised and cancer patients who may not fully benefit from a vaccine. COVID-19 Convalescent plasma (CCP), collected from patients who have recovered from COVID-19, has emerged as a promising therapeutic for COVID-19 patients.

Pre-clinical data on monoclonal antibodies targeting the receptor-binding domain (RBD) of the spike protein of SARS-CoV-2 were promising (1) and clinical studies with neutralizing antibodies targeting SARS-CoV-2 showed some benefit in the outpatient setting, as shown by reductions in viral load and symptom severity (2, 3). However, hospitalized patients did not benefit from monoclonal antibodies targeting the spike protein, and the study was terminated for futility (4).

Preliminary data from small cohort studies have suggested that CCP collected from individuals who have recovered from COVID-19 might be effective for the treatment of patients with severe COVID-19 (5-7). Based on this preliminary evidence, a nationwide study was initiated making CCP available at acute care facilities to individuals with documented SARS-CoV-2 infection and who had severe or life-threatening COVID-19 symptoms, or who were judged by a provider to be at high risk of progression to severe or life-threatening disease. Our sub-cohort study aimed to identify the specific antibodies found in CCP and to estimate the association of their levels with patient survival.

## Methods

### Patient population

This study included adult patients ≥18 years of age who were hospitalized at the MD Anderson Cancer Center, were diagnosed with acute COVID-19 between 4/30/2020 and 8/20/2020, and received one or more doses of CCP as a part of their treatment. This study was part of a nationwide expanded access protocol led by Mayo Clinic investigators for the administration of CCP for the treatment of COVID-19 (NCT04338360). The primary endpoint of the larger study was availability of CCP and the secondary endpoints were safety and serious adverse events related to CCP administration. Inclusion criteria included adult age, laboratory confirmed infection with SARS-CoV-2, and severe or life threatening COVID-19, as judged by the treating physicians to be at high risk of progression to severe or life-threatening disease.

Our sub-study’s primary endpoint was survival time, including survival to discharge, with secondary endpoints being safety. Survival regression models included antibody levels and baseline and cancer-related patient characteristics.

### Antibody testing

The levels of 10 anti-SARS-CoV-2 antibodies (5 IgG isotypes and 5 IgM isotypes) were measured in the CCP products using the Maverick SARS-CoV-2 Multi-Antigen Serology Panel, along with an additional 8 antibodies against antigens of other coronavirus strains (Genalyte, Austin, TX). The 10 anti-SARS-CoV-2 antibodies were measured in each of the CCP products that were administered to all 44 patients in our study, while the 8 additional antibodies against other coronavirus strains were measured in the CCP products of 25 of those patients. This discrepancy in sampling was because the 8 other anti-coronavirus-associated protein antibody tests had not been developed at the time the study commenced. The assay values (given in arbitrary units) are semiquantitative, linear, and represent the reactivity of the patient sample to sample antigens against an established baseline.

### Statistical Methods

Patient characteristics were summarized overall and within the two subgroups [Alive at discharge] (n=32) and [Died during hospitalization] (n=12) using mean (range) for numerical variables and count (%) for categorical variables, with comparisons between the two groups done using Wilcoxon-Mann-Whitney tests or generalized Fisher exact tests. Due to the limited sample size (n=44), a separate Bayesian exponential survival time regression model (8) was fit for each of the 18 antibodies in the CCP product, and also included sex of the patient and an indicator of whether the CCP was administered within 3 days of COVID-19 diagnosis. Patient age was also included in 10 of the 18 models based on preliminary analyses. The exponential distribution was chosen based on better fits compared to a Weibull model, using Deviance Information Criterion statistics (9). In each regression model, to obtain numerical stability, the plasma variable was standardized to a value Z by subtracting the sample mean and dividing by the sample standard deviation. In each model, it was assumed that log (mean survival time) = b_0_ + b_1_ age + b_2_[Female] + b_3_[time to infusion ≤ 3 days] + b_4_Z, with the parameter b_4_ quantifying the additional effect of the antibody level on survival time. Noninformative normal priors with mean = 0 and variance = 10 were assumed for all model parameters. For each antibody, the posterior probability that a larger value had a beneficial effect of on survival time was computed, PBE = (b_4_ > 0 | data). For example, if PBE = 0.95, then the odds are 19 to 1 that a larger antibody value was associated with longer survival time. The probability of a harmful effect is PHE = 1 – PBE. To compare mean antibody value distributions between the two subgroups [Died during hospitalization] (j=1) and [Alive at discharge] (j=2), assuming noninformative normal priors for the means μ_1_ and μ_2_ of Z in the two subgroups, we computed Pr(μ_1 <_ μ_2_ | data) = posterior probability that the mean antibody level was larger for the patients who survived and were discharged than for the patients who died in hospital.

## Results

### Patient and donor characteristics

Our cohort included 44 patients who were hospitalized, diagnosed with acute COVID-19, and received CCP as a part of their treatment. The median age was 60 years (range 37-84) and 38 (86%) had one or more comorbidities. Twenty-five (57%) were male and 19 (43%) were female (**Table 1**). Twenty-eight patients (64%) had active cancer, while 16 patients (36%) were in remission or had pre-malignant disease. The primary diagnosis was a hematologic malignancy in 27 patients (61%), while 17 patients (39%) had a solid tumor. Of the patients with a solid tumor, 6 (38%) had metastatic disease.

**Table 1.**
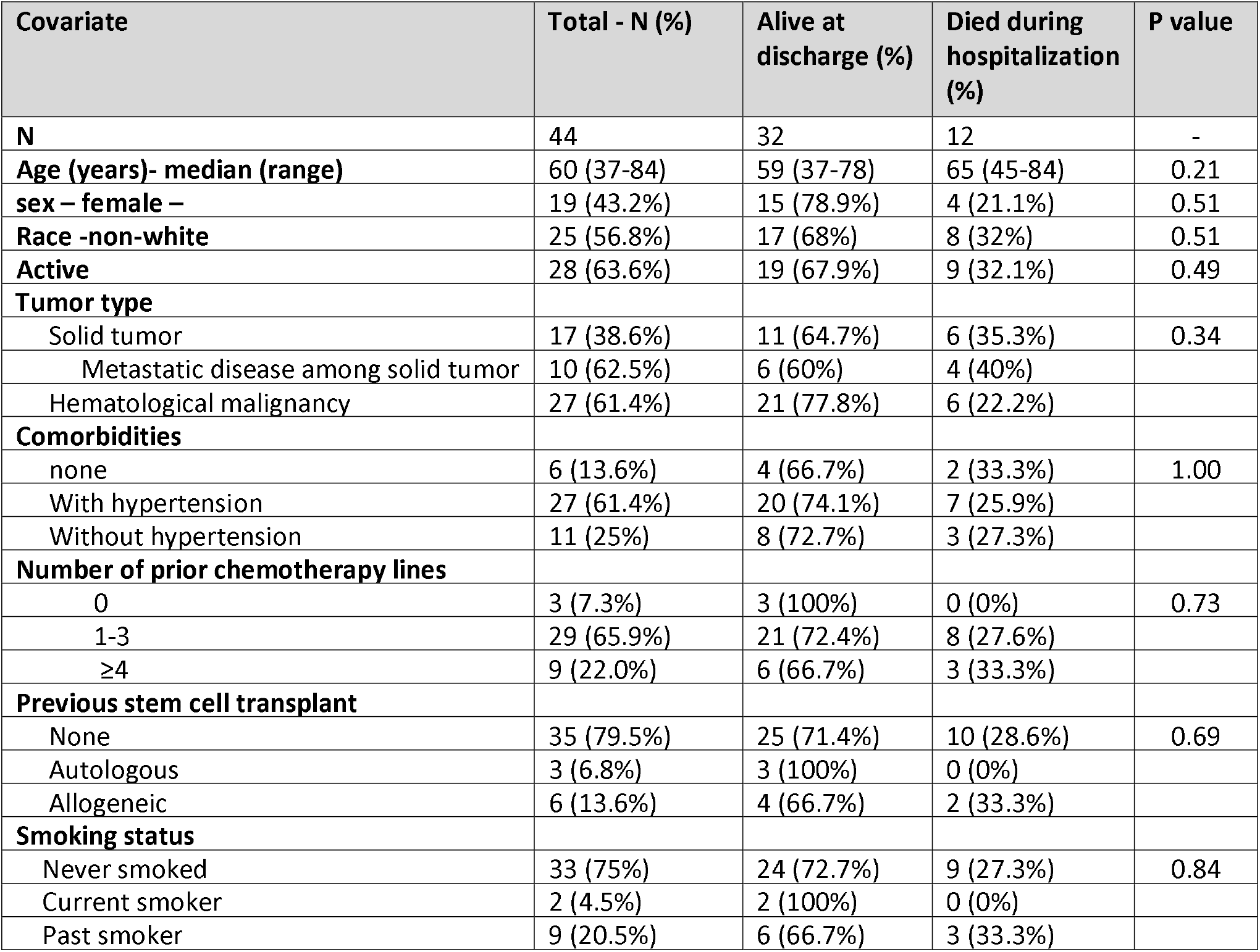
Patient demographic characteristics.

| Covariate | Total - N (%) | Alive at discharge (%) | Died during hospitalization (%) | P value |
| --- | --- | --- | --- | --- |
| **N** | 44 | 32 | 12 | - |
| **Age (years)- median (range)** | 60 (37-84) | 59 (37-78) | 65 (45-84) | 0.21 |
| **sex – female –** | 19 (43.2%) | 15 (78.9%) | 4 (21.1%) | 0.51 |
| **Race -non-white** | 25 (56.8%) | 17 (68%) | 8 (32%) | 0.51 |
| **Active** | 28 (63.6%) | 19 (67.9%) | 9 (32.1%) | 0.49 |
| **Tumor type** |  |  |  |  |
| Solid tumor | 17 (38.6%) | 11 (64.7%) | 6 (35.3%) | 0.34 |
| Metastatic disease among solid tumor | 10 (62.5%) | 6 (60%) | 4 (40%) |  |
| Hematological malignancy | 27 (61.4%) | 21 (77.8%) | 6 (22.2%) |  |
| **Comorbidities** |  |  |  |  |
| none | 6 (13.6%) | 4 (66.7%) | 2 (33.3%) | 1.00 |
| With hypertension | 27 (61.4%) | 20 (74.1%) | 7 (25.9%) |  |
| Without hypertension | 11 (25%) | 8 (72.7%) | 3 (27.3%) |  |
| **Number of prior chemotherapy lines** |  |  |  |  |
| 0 | 3 (7.3%) | 3 (100%) | 0 (0%) | 0.73 |
| 1-3 | 29 (65.9%) | 21 (72.4%) | 8 (27.6%) |  |
| ≥4 | 9 (22.0%) | 6 (66.7%) | 3 (33.3%) |  |
| **Previous stem cell transplant** |  |  |  |  |
| None | 35 (79.5%) | 25 (71.4%) | 10 (28.6%) | 0.69 |
| Autologous | 3 (6.8%) | 3 (100%) | 0 (0%) |  |
| Allogeneic | 6 (13.6%) | 4 (66.7%) | 2 (33.3%) |  |
| **Smoking status** |  |  |  |  |
| Never smoked | 33 (75%) | 24 (72.7%) | 9 (27.3%) | 0.84 |
| Current smoker | 2 (4.5%) | 2 (100%) | 0 (0%) |  |
| Past smoker | 9 (20.5%) | 6 (66.7%) | 3 (33.3%) |  |

Forty-two patients (95%) received one dose of CCP, while 2 patients (5%) received two doses (**Table 2**). The mean unit volume of the CCP products was 240 milliliters. The units were collected from 32 donors who previously had been positive for SARS-CoV-2 infection by either PCR or antibody assays testing for the presence of 5 anti-SARS-CoV-2 antibodies. All donors were asymptomatic for at least 28 days. Time from COVID-19 diagnosis to plasma administration was ≤3 days for 15 patients (34.1%) and 4-7 days for 29 patients (65.9%).

**Table 2-.** COVID-19 related treatment and blood values.

|  | **N (%)** |
| --- | --- |
| **Plasma units administered** |  |
| 1 | 42 (95.5%) |
| 2 | 2 (4.5%) |
| **Other therapies** |  |
| Remdesivir | 32 (72.7%) |
| Tocilizumab | 20 (45.5%) |
| Number of doses |  |
| 0 | 24 (54.5%) |
| 1 | 6 (13.6%) |
| 2 | 12 (27.3%) |
| 3 | 2 (4.5%) |
| Anakinra | 11 (25%) |
| Steroids |  |
| no | 5 (11.4%) |
| yes | 39 (88.6%) |
| Vitamin C | 6 (13.6%) |
| Zinc | 5 (11.4%) |
| **Maximum Oxygen intervention** |  |
| Intubation | 7 (15.9%) |
| High flow oxygen | 14 (31.8%) |
| Nasal cannula | 23 (52.2%) |
| **Maximum blood test values (mean +/- SD)** |  |
| CRP | 150.16 ± 69.03 |
| TNF | 30.07 ± 27.08 |
| IL6 | 294.74 ± 565.24 |
| FERRITIN | 3568.16 ± 5362.36 |
| ANC | 2.67 ± 2.21 |

### COVID-19 management

During hospitalization, 7 patients (16%) required intubation. Five additional patients who had an indication for intubation elected not to be intubated and chose end of life care. Other agents used for COVID-19 treatment in these patients included steroids, zinc, vitamin C, Remdesivir, Tocilizumab, and Anakinra (**Table 2**). Steroids were given based on the dose and schedule described in the RECOVERY trial (10). Remdesivir was given for five days, as previously described (11, 12). Both Anakinra and Tocilizumab were administered to reduce the acute inflammatory responses of COVID-19, as previously described (13-15).

### Safety of CCP administration

Following CCP administration, one patient had a serious adverse event that was deemed to possibly be related to the CCP infusion. The patient needed increased O2 support, which occurred concurrently with the appearance of a rash and generalized itching. This required temporary suspension of the infusion, and supportive care. Shortly thereafter the symptoms subsided, and the patient was able to resume the transfusion and complete the full dose without further adverse events. Twenty-one other patients experienced increased oxygen demand following the CCP administration, which was determined to be unlikely due to the CCP but rather to the underlying compromised respiratory status of the patients. The risk for transfusion-related acute lung injury was mitigated by HLA testing for female donors, and transfusion-associated circulatory overload was mitigated by transfusing the units over 4 hours and meticulous fluid balance follow-up.

### Outcomes and factors predicting survival

Of the 44 patients receiving CCP, 32 survived to discharge (73%) and 12 patients (27%) died during hospitalization. A shorter time from COVID-19 diagnosis to plasma administration, ≤3 days versus >3 days was associated with better survival in males, in which 100% of males with a COVID-19 diagnosis ≤3 days prior to CCP administration survived (95% CI, 54%-100%). In comparison, 58% males who received CCP >3 days after COVID-19 diagnosis survived (95% CI, 34%-80%, long-rank p-value=0.05). However, this was not observed in female patients, in which 86% of females who received CCP ≤3 days after COVID-19 diagnosis survived (95% CI, 42%-100%), compared to 75% who received CCP >3 days after diagnosis survived (95% CI, 42%-95%, long-rank p-value=0.68) (Figure 1).

**Figure 1:**
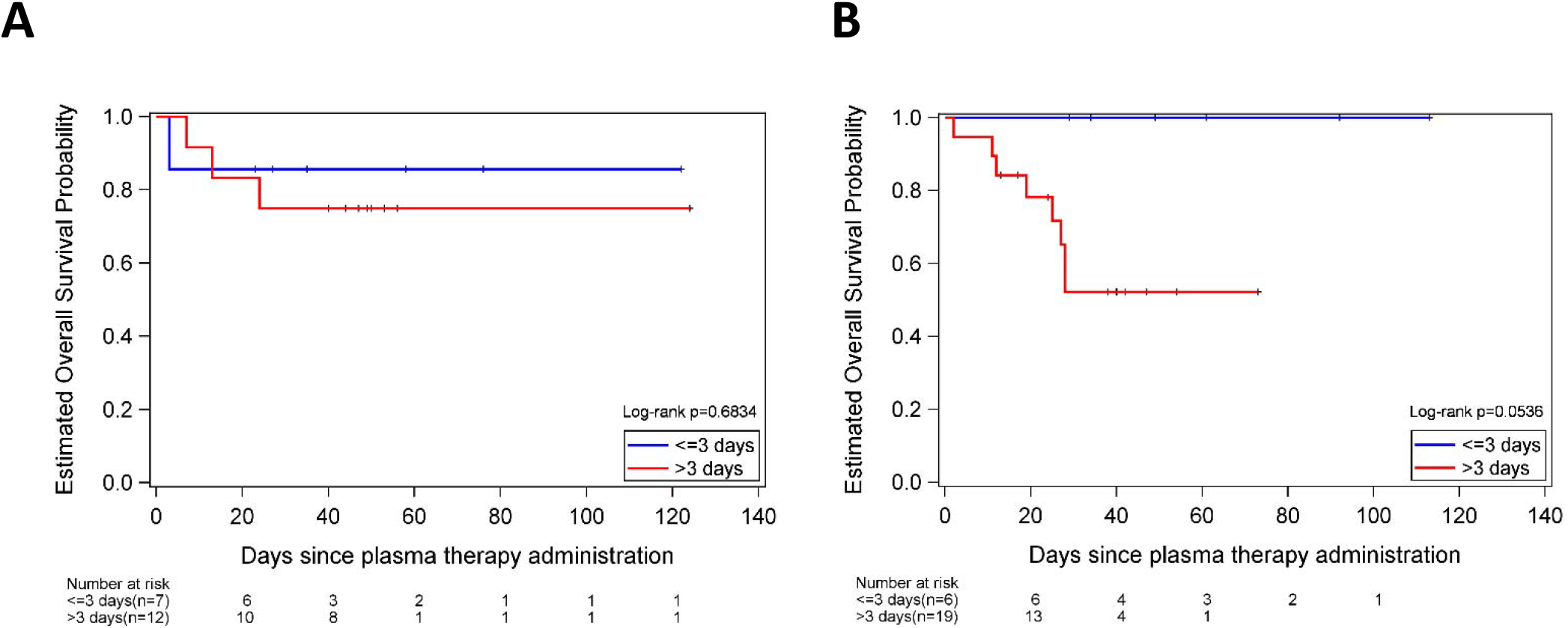
Estimated overall survival probability in females (A) and males (B), stratified by time to plasma administration, ≤3 versus >3 days of COVID-19 diagnosis.

We compared levels of antibodies against SARS-CoV-2 antigens and against antigens of other coronavirus strains in the CCPs administered to patients who survived and died after receiving CCP. Interestingly, we found that the largest differences in antibody levels between patients who survived compared to those who died were of IgGs against the spike proteins of two non-SARS-CoV-2 coronavirus strains, HCoV-OC43 and HCoV-HKU1 (Figure 2).

**Figure 2:**
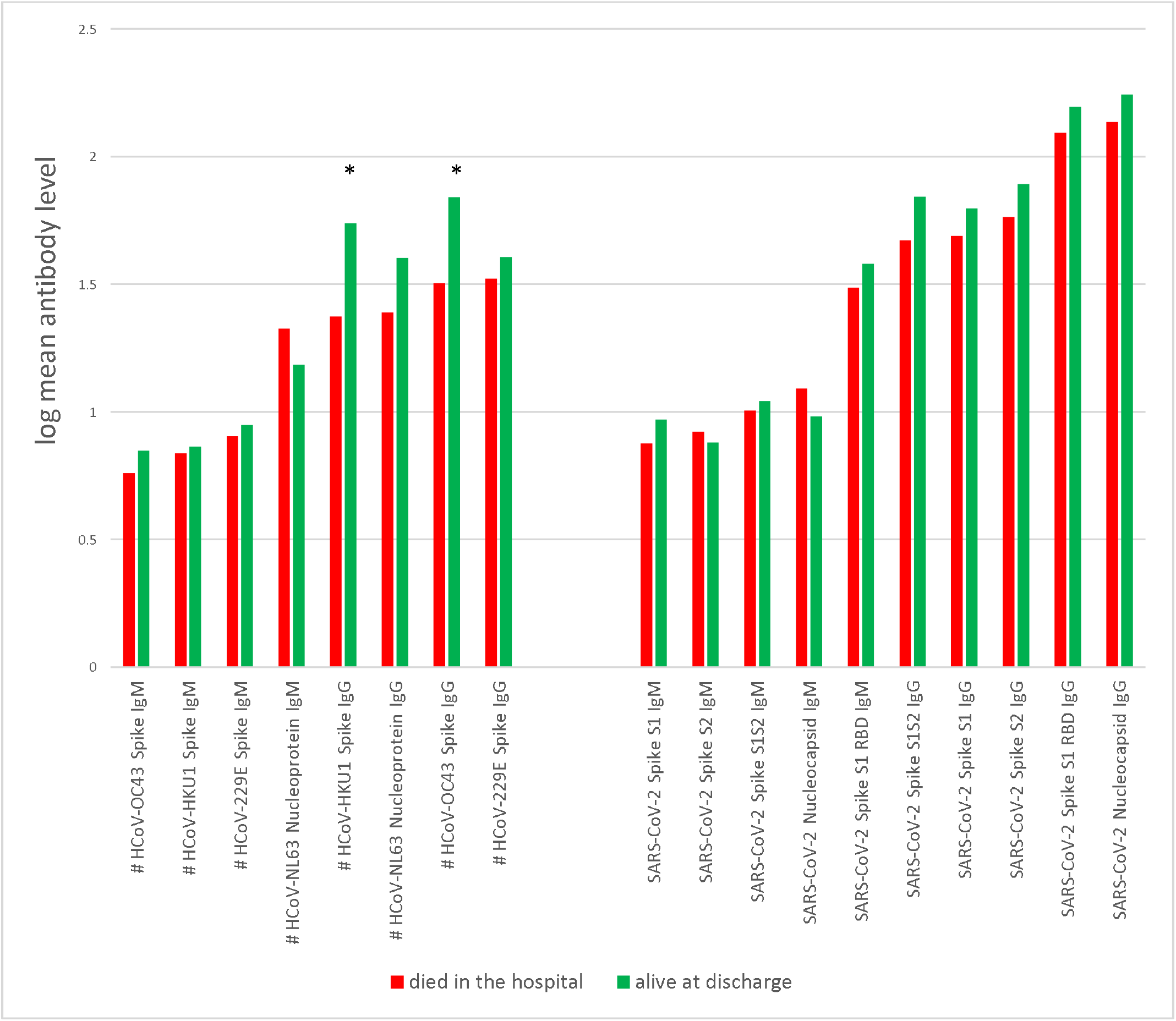
Mean levels for each antibody, for patients who died in hospital (red) and survived to discharge (green). *, probability of beneficial effect = 1; #, data available for 25 of 44 patients.

Figure 3 and Supplemental Table 1 summarize PBE values for the 18 antibodies tested, computed while accounting for age (when significant in a univariate analysis), sex, and time to CCP administration. Prognostically, younger age (PBE range = 0.77 to 0.88 in the 18 fitted models), female sex (PBE range = 0.81 to 0.97), and plasma administration ≤3 days from COVID-19 diagnosis (PBE range = 0.84 to 0.98) were associated with longer survival. For each of the two IgG antibodies targeting HCoV-OC43 spike and HCoV-HKU1 spike, the PBE was 1.00. Three SARS-CoV-2 specific antibodies had a PBE of 0.90 to 0.95. Four antibodies, all of IgM isotype, had PBEs less than 0.80.

**Figure 3:**
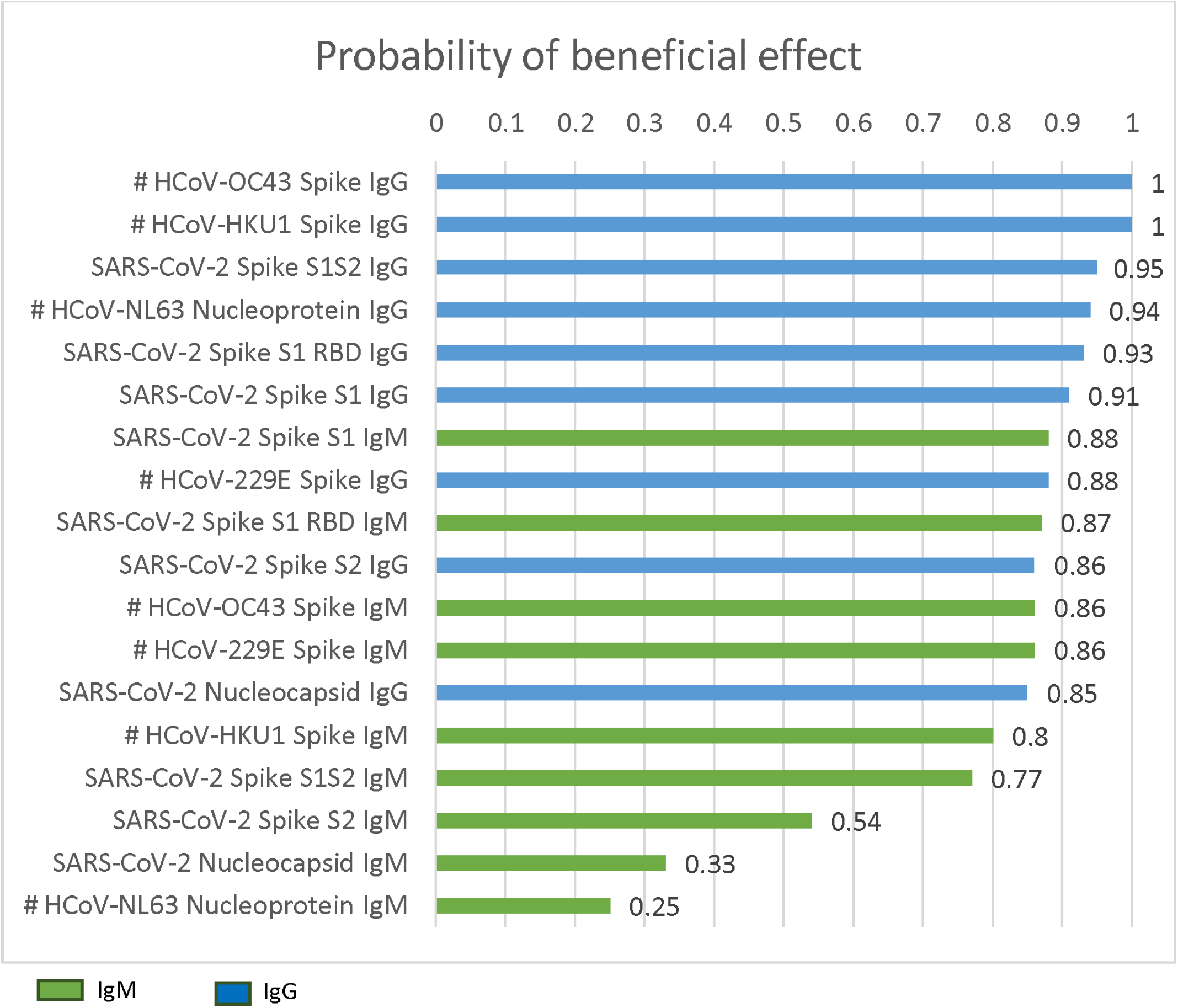
Posterior probability of beneficial effect of a larger value of each antibody, accounting for age, sex, and time to CCP administration. #, data available for 25 of 44 patients.

Furthermore, as shown in Figure 4, we found that the probability of survival to discharge was correlated with the amount of IgG targeting HCoV-OC43 spike protein in the CCP, after adjusting for sex and time to CCP administration. This trend was seen in both male and female patients, as well as those who received CCP ≤3 days from COVID-19 diagnosis and those who received CCP >3 days from COVID-19 diagnosis. Similar findings were found for HCoV-HKU1(data not shown).

**Figure 4:**
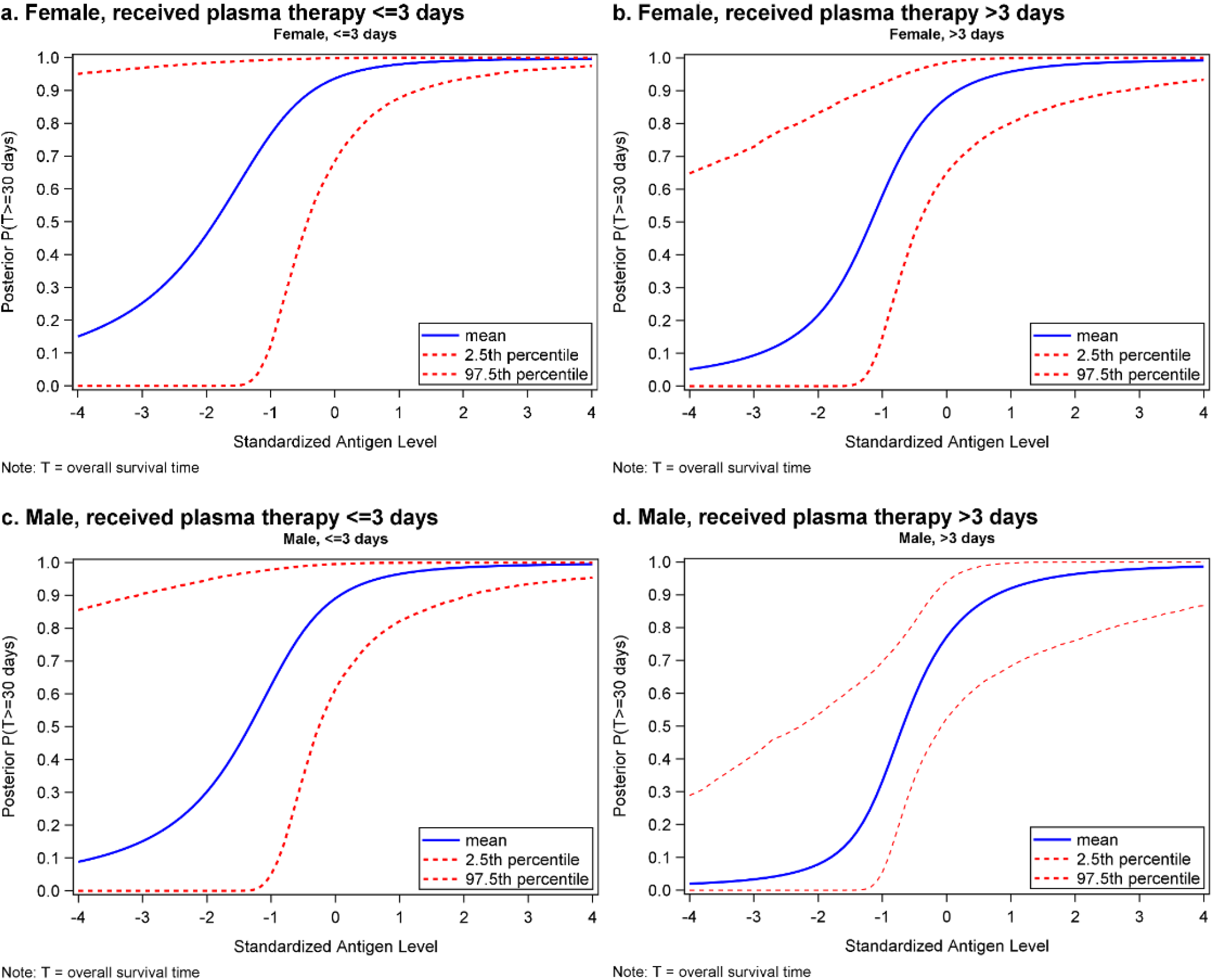
Posterior probability of surviving at least 30 days as a function of standardized HCoV-OC43 spike IgG antibody level, by sex and time to plasma administration in females who received CCP ≤3 days (**A**), females who received CCP >3 days (**B**), males who received CCP ≤3 days (**C**), and males who received CCP >3 days (**D**) from COVID-19 diagnosis.

## Discussion

The aim of our study was to identify which CCP components may confer a survival benefit for patients with moderate to severe COVID-19 and quantify such benefits. Interestingly, we found that higher levels of two IgG antibodies against coronavirus strains besides SARS-CoV-2, which targeted HCoV-OC43 spike and HCoV-HKU1 spike proteins, were associated with a larger probability of being discharged alive from the hospital. In separate regression analyses, the PBE was 1.00 for both antibodies, after adjusting for age, sex, and time from COVID-19 diagnosis to CCP administration.

Larger studies have examined the effect of CCP on clinical outcomes of COVID-19 patients. Our cohort was part of a large study initiated at the Mayo Clinic that recently reported on safety, and similar to our findings, did not find severe toxicity associated with CCP (16). The mortality rate within 4 hours of plasma infusion was 0.3% (63 cases of 20,000 patients), and in our trial no deaths were recorded within this timeframe. Salazar et al. performed a prospective, propensity score matched study looking at mortality within 28 days of receiving CCP in COVID-19 patients and a matched control cohort who did not receive the plasma. They showed a reduction in mortality following CCP administration (17). Similar to our findings, this improvement was seen particularly in the patients transfused within 3 days of admission, and those with a higher anti-SARS-CoV-2 spike protein antibody titer. The recently published report of the Mayo Clinic study, which included 3,082 patients transfused with CCP, showed a reduction in mortality for patients transfused within 3 days of COVID-19 diagnosis (18).

Both of these trials evaluated the effect of CCP on large patient populations, but the specific components of CCP tested differ between them. The Mayo Clinic study used a qualitative assay (the Ortho-Clinical Diagnostics VITROS Anti-SARS-CoV-2 IgG chemiluminescent immunoassay) testing for the spike subunit 1 (S1) protein and showed that higher levels of this antibody correlated with improved survival. Salazar et al. evaluated antibodies aimed at the ectodomain and RBD components of the SARS-CoV-2 spike protein and showed a reduction in mortality that correlated with increasing anti-SARS-CoV-2 spike protein antibody titers. In our study, using the Maverick SARS-CoV-2 Multi-Antigen Serology Panel, we evaluated levels of antibodies against SARS-CoV-2 proteins and determined their clinical benefit. We found that SARS-CoV-2 spike S1S2 IgG, SARS-CoV-2 spike S1 RBD IgG and SARS-CoV-2 spike S1 IgG had PBEs of 0.95, 0.93 and 0.91, respectively, which indicates that these antibodies have a clinical benefit. However, somewhat surprisingly and possibly due to the small sample size of 25 patients in whom those two antibodies were measured, the 2 antibodies with the highest PBE = 1.00 were those targeting the HCoV-OC43 and HCoV-HKU1 spike proteins, which are found in previously reported strains of coronaviruses (19, 20).

SARS-CoV-2 has two known targets used to attach to the host’s cells and begin the viral replication cycle. The most extensively studied is the angiotensin-converting enzyme 2 (ACE2) receptor, to which the spike S1 RBD binds (21, 22). Monoclonal antibodies targeting this domain are currently under investigation as a possible therapeutic. Another point of attachment, which is also common to other coronavirus strains including HCoV-OC43 and HCoV-HKU1, is the ganglioside-rich domain (GRD) binding site on the cell’s plasma membrane, using the viral N-terminal domain (23, 24).

A study looking at the binding of HCoV-OC43 and HCoV-HKU1 found a conserved 9-O-acetylated sialic acid receptor-binding site as a common site for both strains, as part of a GRD binding site within the spike protein (25). While our findings need further verification, antibodies directed at the 9-O-acetylated sialic acid receptor-binding site, or other sites common to SARS-CoV-2 and other coronavirus strains, may be an improved target for SARS-CoV-2 neutralization (26). This hypothesis is supported by our finding that these antibodies had the highest PBEs of all antibodies tested, including the commonly studied SARS-CoV-2 S1 spike and RBD IgGs.

Some evidence exists as to the protective effect of prior common coronavirus infections on the clinical course of COVID-19. A recent trial looking at patients with a respiratory PCR panel positive for one of 4 common coronavirus strains, including HCoV-OC43 and HCoV-HKU1, and were then hospitalized with COVID-19, showed marked reduction in intensive care utilization and mechanical ventilation, signifying a milder disease course (27). There is evidence that patients may have prior T cell immunogenicity against SARS-CoV-2, even without a documented infection, which indicates T cell cross-reactivity between the different strains (28, 29). However, to the best of our knowledge, this is the first evidence that the transfer of antibodies common to both SARS-CoV-2 and other coronaviruses confers clinical benefit for COVID-19 patients. This is especially important, as recent studies have not shown benefit for monoclonal antibodies targeting the SARS-CoV-2 spike protein for hospitalized patients (4) and other targets may confer better responses in patients with more severe disease. The conserved 9-O-acetylated sialic acid receptor-binding site may be an alternative target to other regions of the spike protein, which have demonstrated rapidly developing mutations that may render previous immunogenicity less effective (30, 31).

Our cohort size was small and did not include a randomized control group. Thus, we could not calculate the relative effect of each antibody compared to the others on the panel. A larger cohort is needed to assess whether there is a correlation between the different antibodies and if they may work synergistically together. However, our findings may assist in the development of antibodies directed at sites common to both SARS-CoV-2 and other coronaviruses, paving the way for clinical intervention trials evaluating effects of anti-HCoV-OC43 and HCoV-HKU1 antibodies in COVID-19 patients. This is especially relevant, since previous studies have shown that using a combination of antibodies may reduce the likelihood of mutations that allow for viral resistance (1). Even if a vaccine proves to be an effective preventative measure, it is likely that a certain proportion of the population, particularly immunocompromised cancer patients, may not be able to mount a robust immune reaction and may need passive antibody administration to treat this serious infection.

In conclusion, our study evaluated the levels of 18 different antibodies found in patients who recovered from COVID-19, and found anti-HCoV-OC43 and HCoV-HKU1 IgG antibodies to have the highest estimated beneficial value, particularly if given within 3 days of COVID-19 diagnosis. These antibodies, possibly targeting a different domain than those studied thus far, provide a new, promising therapeutic target for future monoclonal antibody production. Larger interventional studies are needed to confirm their clinical benefit and determine if there is a synergistic effect with other antibodies.

## Data Availability

Please contact the corresponding author for study dataset availability

## Author contributions

*UG, JLR, and EJS contributed collecting and verifying the underlying data, as well as with drafting the paper, and writing the manuscript; KK, FM, and CK participated in generating the patient safety data as well as the antibody testing; JS and PFT participated in analyzing the data; All other authors participated in active follow-up and reporting on patient outcomes, as well as in the critical revision of the article; all authors reviewed and approved the manuscript*.

## Acknowledgments

The statistical analysis work was supported in part by the Cancer Center Support Grant (NCI Grant P30 CA016672).

Dr. Greenbaum is a recipient of a fellowship Grant from the American Physicians’ Fellowship for Medicine in Israel (APF).

## Disclaimer

The views and opinions expressed in this publication are those of the authors and do not reflect the official policy or position of the US Department of Health and Human services and its agencies including the Biomedical Research and Development Authority and the Food and Drug Administration, as well as any agency of the U.S. government. Assumptions made within and interpretations from the analysis are not reflective of the position of any U.S. government entity.

## References

1. Baum A, Fulton BO, Wloga E, Copin R, Pascal KE, Russo V, et al. Antibody cocktail to SARS-CoV-2 spike protein prevents rapid mutational escape seen with individual antibodies. Science. 2020;369(6506):1014–8.

2. Chen P, Nirula A, Heller B, Gottlieb RL, Boscia J, Morris J, et al. SARS-CoV-2 Neutralizing Antibody LY-CoV555 in Outpatients with Covid-19. N Engl J Med. 2020;384(3):229–37.

3. Weinreich DM, Sivapalasingam S, Norton T, Ali S, Gao H, Bhore R, et al. REGN-COV2, a Neutralizing Antibody Cocktail, in Outpatients with Covid-19. N Engl J Med. 2021;384(3):238–51.

4. Group A-TL-CS, Lundgren J, Grund B, Barkauskas C, Holland T, Gottlieb R, et al. A Neutralizing Monoclonal Antibody for Hospitalized Patients with Covid-19. N Engl J Med. 2020;Epub ahead of print.

5. Duan K, Liu B, Li C, Zhang H, Yu T, Qu J, et al. Effectiveness of convalescent plasma therapy in severe COVID-19 patients. Proc Natl Acad Sci USA. 2020;117(17):9490–6.

6. Shen C, Wang Z, Zhao F, Yang Y, Li J, Yuan J, et al. Treatment of 5 critically ill patients with COVID-19 with convalescent plasma. JAMA. 2020;323(16):1582–9.

7. Li L, Zhang W, Hu Y, Tong X, Zheng S, Yang J, et al. Effect of Convalescent Plasma Therapy on Time to Clinical Improvement in Patients With Severe and Life-threatening COVID-19: A Randomized Clinical Trial. JAMA. 2020;324(5):460–70.

8. Ibrahim J, Chen M, Sinha D. Bayesian methods for joint modeling of longitudinal and survival data with applications to cancer vaccine trials. Stat Sin. 2004;14(3):863–83.

9. Spiegelhalter DJ, Best NG, Carlin BP, Van Der Linde A. Bayesian measures of model complexity and fit. JR Statistic Soc B. 2002;64(4):583–639.

10. Group RC, Horby P, Lim WS, Emberson JR, Mafham M, Bell JL, et al. Dexamethasone in Hospitalized Patients with Covid-19-Preliminary Report. N Engl J Med. 2020;384(8):693–704.

11. Goldman JD, Lye DC, Hui DS, Marks KM, Bruno R, Montejano R, et al. Remdesivir for 5 or 10 days in patients with severe Covid-19. N Engl J Med. 2020;383(19):1827–37.

12. Wang Y, Zhang D, Du G, Du R, Zhao J, Jin Y, et al. Remdesivir in adults with severe COVID-19: a randomised, double-blind, placebo-controlled, multicentre trial. Lancet. 2020;395(10236):1569–78.

13. Cavalli G, De Luca G, Campochiaro C, Della-Torre E, Ripa M, Canetti D, et al. Interleukin-1 blockade with high-dose anakinra in patients with COVID-19, acute respiratory distress syndrome, and hyperinflammation: a retrospective cohort study. Lancet Rheumatol. 2020;2(6):e325–e31.

14. Xu X, Han M, Li T, Sun W, Wang D, Fu B, et al. Effective treatment of severe COVID-19 patients with tocilizumab. Proc Natl Acad Sci USA. 2020;117(20):10970–5.

15. Guaraldi G, Meschiari M, Cozzi-Lepri A, Milic J, Tonelli R, Menozzi M, et al. Tocilizumab in patients with severe COVID-19: a retrospective cohort study. Lancet Rheumatol. 2020;2(8):e474–e84.

16. Joyner MJ, Bruno KA, Klassen SA, Kunze KL, Johnson PW, Lesser ER, et al. Safety update: COVID-19 convalescent plasma in 20,000 hospitalized patients. Mayo Clin Proc. 2020;95(9):1888–97.

17. Salazar E, Christensen PA, Graviss EA, Nguyen DT, Castillo B, Chen J, et al. Treatment of COVID-19 Patients with Convalescent Plasma Reveals a Signal of Significantly Decreased Mortality. Am J Pathol. 2020;190(11):2290–303.

18. Joyner MJ, Carter RE, Senefeld JW, Klassen SA, Mills JR, Johnson PW, et al. Convalescent Plasma Antibody Levels and the Risk of Death from Covid-19. N Engl J Med. 2021;Epub ahead of print.

19. Woo PCY, Lau SKP, Chu C-m, Chan K-h, Tsoi H-w, Huang Y, et al. Characterization and Complete Genome Sequence of a Novel Coronavirus, Coronavirus HKU1, from Patients with Pneumonia. J Virol. 2005;79(2):884–95.

20. Mounir S, Talbot PJ. Molecular characterization of the S protein gene of human coronavirus OC43. J Gen Virol. 1993;74(9):1981–7.

21. Clausen TM, Sandoval DR, Spliid CB, Pihl J, Perrett HR, Painter CD, et al. SARS-CoV-2 Infection Depends on Cellular Heparan Sulfate and ACE2. Cell. 2020;183(4):1043–57.

22. Wec AZ, Wrapp D, Herbert AS, Maurer DP, Haslwanter D, Sakharkar M, et al. Broad neutralization of SARS-related viruses by human monoclonal antibodies. Science. 2020;369(6504):731–6.

23. Fantini J, Di Scala C, Chahinian H, Yahi N. Structural and molecular modelling studies reveal a new mechanism of action of chloroquine and hydroxychloroquine against SARS-CoV-2 infection. Int J Antimicrob Agents. 2020;55(5):105960.

24. Tortorici MA, Walls AC, Lang Y, Wang C, Li Z, Koerhuis D, et al. Structural basis for human coronavirus attachment to sialic acid receptors. Nat Struct Mol Biol. 2019;26(6):481–9.

25. Hulswit RJG, Lang Y, Bakkers MJG, Li W, Li Z, Schouten A, et al. Human coronaviruses OC43 and HKU1 bind to 9-O-acetylated sialic acids via a conserved receptor-binding site in spike protein domain A. Proc Natl Acad Sci USA. 2019;116(7):2681–90.

26. Morniroli D, Giannì ML, Consales A, Pietrasanta C, Mosca F. Human Sialome and Coronavirus Disease-2019 (COVID-19) Pandemic: An Understated Correlation? Front Immunol. 2020;11:1480.

27. Sagar M, Reifler K, Rossi M, Miller NS, Sinha P, White L, et al. Recent endemic coronavirus infection is associated with less severe COVID-19. J Clin Invest. 2020;131(1):e143380.

28. Le Bert N, Tan AT, Kunasegaran K, Tham CYL, Hafezi M, Chia A, et al. SARS-CoV-2-specific T cell immunity in cases of COVID-19 and SARS, and uninfected controls. Nature. 2020;584(7821):457–62.

29. Mateus J, Grifoni A, Tarke A, Sidney J, Ramirez SI, Dan JM, et al. Selective and cross-reactive SARS-CoV-2 T cell epitopes in unexposed humans. Science. 2020;370(6512):89–94.

30. Volz E, Hill V, McCrone JT, Price A, Jorgensen D, O’Toole Á, et al. Evaluating the Effects of SARS-CoV-2 Spike Mutation D614G on Transmissibility and Pathogenicity. Cell. 2021;184(1):64–75.

31. Andrew Rambaut NL, Oliver Pybus, Wendy Barclay, Jeff Barrett, Alesandro Carabelli, Tom Conno7, Tom Peacock, David L Robertson, Erik Volz. Preliminary genomic characterisation of an emergent SARS-CoV-2 lineage in the UK defined by a novel set of spike mutations 2020 [Available from: https://virological.org/t/preliminary-genomic-characterisation-of-an-emergent-sars-cov-2-lineage-in-the-uk-defined-by-a-novel-set-of-spike-mutations/563.

