## Supplemental Table 1 for "High levels of common cold coronavirus antibodies in convalescent plasma are associated with improved survival in COVID-19 patients"

**Supplemental Table 1**: Bayesian multivariate models for each of the 18 antibodies found in CCP, calculating the probability of a beneficial effect (PBE) on survival to discharge, after accounting for sex, time to administration and age (when significant in the univariate analysis).

| Model |  |  | Posterior Quantities | | | | |
| --- | --- | --- | --- | --- | --- | --- | --- |
|  | Antibody | Covariate | Posterior mean | 95% credible interval | | | PBE P(beta>0) |
| 1 | HCoV-OC43 Spike IgG^2^ | Female | 0.8395 | -0.6923 | 2.681 | 0.85 | |
|  |  | Administered >3 days | -1.2221 | -4.0632 | 0.8819 | 0.16 | |
|  |  | Standardized antibody level | 1.5583 | 0.2923 | 3.1043 | **1.00** | |
| 2 | HCoV-HKU1 Spike IgG^2^ | Female | 1.0331 | -0.4747 | 2.8861 | 0.90 | |
|  |  | Administered >3 days | -1.3685 | -4.0935 | 0.6992 | 0.12 | |
|  |  | Standardized antibody level | 1.496 | 0.2449 | 3.3112 | **1.00** | |
| 3 | SARS-CoV-2 Spike S1S2 IgG^1^ | Age | -0.0298 | -0.0809 | 0.0193 | 0.12 | |
|  |  | Female | 1.2522 | -0.039 | 2.6107 | 0.97 | |
|  |  | Administered >3 days | -1.1937 | -3.0538 | 0.2323 | 0.06 | |
|  |  | Standardized antibody level | 0.7403 | -0.1225 | 1.8117 | **0.95** | |
| 4 | HCoV-NL63 Nucleoprotein IgG^2^ | Female | 0.8307 | -0.6918 | 2.6705 | 0.84 | |
|  |  | Administered >3 days | -1.7137 | -4.3618 | 0.3093 | 0.05 | |
|  |  | Standardized antibody level | 1.0178 | -0.1236 | 2.9143 | **0.94** | |
| 5 | SARS-CoV-2 Spike S1 RBD IgG^1^ | Age | -0.0268 | -0.078 | 0.0229 | 0.15 | |
|  |  | Female | 1.3019 | -0.0438 | 2.7873 | 0.97 | |
|  |  | Administered >3 days | -1.3706 | -3.1735 | 0.0821 | 0.03 | |
|  |  | Standardized antibody level | 0.4967 | -0.1479 | 1.2625 | **0.93** | |
| 6 | SARS-CoV-2 Spike S1 IgG^1^ | Age | -0.0305 | -0.0833 | 0.0209 | 0.12 | |
|  |  | Female | 1.2838 | -0.1033 | 2.7935 | 0.96 | |
|  |  | Administered >3 days | -1.4639 | -3.257 | -0.0348 | 0.02 | |
|  |  | Standardized antibody level | 0.4126 | -0.1793 | 1.0826 | **0.91** | |
| 7 | SARS-CoV-2 Spike S1 IgM^1^ | Age | -0.0268 | -0.0788 | 0.0242 | 0.15 | |
|  |  | Female | 1.1339 | -0.2172 | 2.5965 | 0.95 | |
|  |  | Administered >3 days | -1.4761 | -3.2261 | -0.0719 | 0.02 | |
|  |  | Standardized antibody level | 0.3611 | -0.228 | 1.004 | 0.88 | |
| 8 | HCoV-229E Spike IgG^2^ | Female | 0.8763 | -0.6155 | 2.7198 | 0.86 | |
|  |  | Administered >3 days | -1.7867 | -4.4813 | 0.212 | 0.04 | |
|  |  | Standardized antibody level | 0.4766 | -0.2697 | 1.3887 | 0.88 | |
| 9 | SARS-CoV-2 Spike S1 RBD IgM^1^ | Age | -0.0225 | -0.0702 | 0.0235 | 0.17 | |
|  |  | Female | 0.9581 | -0.2486 | 2.277 | 0.94 | |
|  |  | Administered >3 days | -1.3575 | -3.159 | 0.1209 | 0.04 | |
|  |  | Standardized antibody level | 0.4076 | -0.2546 | 1.18 | 0.87 | |
| 10 | SARS-CoV-2 Spike S2 IgG^1^ | Age | -0.0279 | -0.0793 | 0.0215 | 0.13 | |
|  |  | Female | 1.1785 | -0.1565 | 2.5976 | 0.95 | |
|  |  | Administered >3 days | -1.3863 | -3.1524 | 0.0547 | 0.03 | |
|  |  | Standardized antibody level | 0.4896 | -0.2952 | 1.454 | 0.86 | |
| 11 | HCoV-OC43 Spike IgM^2^ | Female | 1.0869 | -0.4563 | 2.9297 | 0.90 | |
|  |  | Administered >3 days | -1.8244 | -4.4395 | 0.1881 | 0.04 | |
|  |  | Standardized antibody level | 0.3682 | -0.3046 | 1.0813 | 0.86 | |
| 12 | HCoV-229E Spike IgM^2^ | Female | 1.2289 | -0.4112 | 3.1422 | 0.92 | |
|  |  | Administered >3 days | -1.7941 | -4.4919 | 0.2499 | 0.05 | |
|  |  | Standardized antibody level | 0.4483 | -0.3646 | 1.361 | 0.86 | |
| 13 | SARS-CoV-2  Nucleocapsid IgG^1^ | Age | -0.0256 | -0.0761 | 0.0234 | 0.15 | |
|  |  | Female | 1.1347 | -0.1848 | 2.5785 | 0.95 | |
|  |  | Administered >3 days | -1.4729 | -3.3068 | -0.0437 | 0.02 | |
|  |  | Standardized antibody level | 0.3265 | -0.2642 | 1.0121 | 0.85 | |
| 14 | HCoV-HKU1 Spike IgM^2^ | Female | 1.1411 | -0.4932 | 3.103 | 0.90 | |
|  |  | Administered >3 days | -1.7647 | -4.4361 | 0.2983 | 0.05 | |
|  |  | Standardized antibody level | 0.3082 | -0.4041 | 1.0585 | 0.80 | |
| 15 | SARS-CoV-2  Spike S1S2 IgM^1^ | Age | -0.0217 | -0.07 | 0.0259 | 0.19 | |
|  |  | Female | 0.986 | -0.2624 | 2.3081 | 0.94 | |
|  |  | Administered >3 days | -1.374 | -3.253 | 0.1334 | 0.04 | |
|  |  | Standardized antibody level | 0.2526 | -0.3745 | 0.9837 | 0.77 | |
| 16 | SARS-CoV-2  Spike S2 IgM^1^ | Age | -0.0204 | -0.0692 | 0.0276 | 0.20 | |
|  |  | Female | 0.9194 | -0.3289 | 2.2747 | 0.92 | |
|  |  | Administered >3 days | -1.4734 | -3.2789 | -0.0046 | 0.02 | |
|  |  | Standardized antibody level | 0.0456 | -0.4307 | 0.6267 | 0.54 | |
| 17 | SARS-CoV-2  Nucleocapsid IgM^1^ | Age | -0.0183 | -0.0677 | 0.0294 | 0.23 | |
|  |  | Female | 0.8822 | -0.3145 | 2.2222 | 0.92 | |
|  |  | Administered >3 days | -1.4416 | -3.2611 | 0.0179 | 0.03 | |
|  |  | Standardized antibody level | -0.0939 | -0.5375 | 0.4285 | 0.33 | |
| 18 | HCoV-NL63 NucleoproteinIgM^2^ | Female | 0.7347 | -0.8214 | 2.5691 | 0.81 | |
|  |  | Administered >3 days | -1.6808 | -4.3567 | 0.3677 | 0.06 | |
|  |  | Standardized antibody level | -0.1728 | -0.659 | 0.4447 | 0.25 | |

^1^ age, sex, and time to administration were adjusted

^2^ sex and time to administration were adjusted
